# Patterns of growth in childhood in relation to adult schooling attainment and IQ in 6 birth cohorts in low and middle-income countries: evidence from COHORTS

**DOI:** 10.1101/2020.06.16.20133074

**Authors:** Natalia E Poveda, Fernando P Hartwig, Cesar G Victora, Linda S Adair, Fernando C Barros, Santosh K Bhargava, Bernardo L Horta, Nanette R Lee, Reynaldo Martorell, Mónica Mazariegos, Ana M B Menezes, Shane A Norris, Linda M Richter, Harshpal Singh Sachdev, Alan Stein, Fernando C Wehrmeister, Aryeh D Stein, COHORTS Group

## Abstract

**Background:** Growth faltering has been associated with poor intellectual performance. The relative strengths of associations between growth in early and in later childhood remain underexplored. We examined the association between growth in childhood and adolescence and adult human capital in five low- or middle-income countries (LMICs).

**Methods:** We analyzed data from six prospective birth cohorts of five LMICs (Brazil, Guatemala, India, the Philippines, and South Africa). We assessed the associations of measures of height and relative weight at four ages (birth, at around age 2 years, mid-childhood (MC), adulthood), with two dimension of adult human capital (schooling attainment and IQ).

**Findings:** In site- and sex-pooled analyses, size at birth and linear growth from birth to around 2 years of age were positively associated with schooling attainment and adult IQ. Linear growth from age 2 years to MC and from MC to adulthood was not associated with higher school attainment or IQ. Change in relative weight in early childhood was not associated with either outcome. Relative weight in MC and in adulthood were inversely associated with schooling attainment but were not associated with adult IQ.

**Interpretation:** Linear growth in the first 1,000 days is a predictor of schooling attainment and IQ in adulthood in LMICs. Linear growth in later periods was not associated with either of these outcomes. Changes in relative weight had inconsistent association with schooling and IQ in adulthood.

**Funding:** Bill and Melinda Gates Foundation (OPP1164115)

**Research in context:** *Evidence before this study:* Early life growth faltering has been associated with poor cognitive and intellectual performance in childhood and poorer schooling outcomes in children and adults. There is a paucity of data about how growth in specific age intervals over the course of childhood and adolescence relates to attained schooling and adult cognitive performance. We conducted a literature search using the terms (growth [Title/Abstract]) AND ((school [Title/Abstract] OR schooling [Title/Abstract]) AND (intelligence [Title/Abstract] OR IQ [Title/Abstract]) OR (human capital [Title/Abstract]) in Pubmed. The search yielded 536 publications from 1965 to 2020. We screened titles and selected 31 publications that included linear growth and our outcomes of interest, namely school attainment and intelligence quotient (IQ). Additionally, we checked reference lists of selected articles and identified eleven papers that were not displayed in the initial electronic query. We therefore reviewed 42 abstracts and identified 24 unique studies conducted in low and middle-income countries (LMICs). Fourteen of them investigated the association of birth size and/or early-life size with schooling or IQ, or with both outcomes during childhood. Three studies investigated the association between linear growth in early childhood and schooling and intelligence in adults, one studied the association between early-life undernutrition and IQ in early adulthood and another six publications investigated the association between growth and school attainment in adults. Economists have also studied the relationship between stunting or linear growth and schooling in LMICs, but to our knowledge not the relative importance of growth during specific age intervals.

*Added value of this study:* This is an analysis of the associations between child and adolescent growth and two dimensions of human capital (schooling attainment and IQ) in adulthood in six birth cohorts from five LMICs. The evidence of long-term associations of linear growth with adult IQ is scarce and the few published studies have analyzed data from a single country. In the present study, we found that size at birth and linear growth from birth to around 2 years of age were positively associated with both schooling and IQ in adulthood. Linear growth between early and mid-childhood (MC)was not associated with higher school attainment or IQ in adjusted models. Linear growth from MC to adulthood was not associated with IQ in men or women, and was inversely associated with schooling attainment in women only. Change in relative weight in early childhood was positively associated with schooling attainment only in minimally adjusted models. Relative weight measures in MC and adulthood were inversely associated with schooling attainment. Change in relative weight between MC and adulthood was not associated with adult IQ.

*Implications of all the available evidence:* We confirmed in multiple cohorts that birth size and linear growth from birth to age 2 years are predictors of schooling attainment and adult IQ. Linear growth in early life was the strongest predictor of these two human capital dimensions in adulthood among individuals in LMICs. We did not find evidence that supports the notion that linear growth in adolescence contributes to a better cognitive performance in adulthood. Thus, our results inform the more effective timing of nutritional and other interventions to improve linear growth and human capital in the long-term.

## Introduction

Growth faltering remains widespread in low and middle-income countries (LMIC), with 96.8million children under five being stunted. ^1^ The period between conception and the second birthday is critical for human growth^2^ as well as for development of brain structure, architecture and function.^3^ In early childhood, sensory and motor functions develop, followed by acquisition of basic language skills and spatial attention.^3^ Evidence from birth cohort studies indicates that linear growth in the first 24 months after birth is among the strongest predictors of school attainment,^4^ being positively associated with more years of schooling, lower probability of failing a grade, younger age of enrollment in school, and better verbal and nonverbal skills.^4-8^Further, lower schooling attainment is related to future disadvantages such as lower wages^7,9^,less income,4 unemployment or informal sector work,^10^ and a higher probability of living in poverty.^7^

Prenatal and postnatal growth predict childhood and adolescent cognition. A meta-analysis of 68 studies from LMIC demonstrated associations between linear growth in children younger than 24 months and cognition and motor skills at 5-11 years of age.^11^ In the multi-country Young Lives Study, growth from 1 to 8 years of age was associated with better performance in mathematics, reading, and vocabulary tests in children aged 8 years.^12^ They also found that stunted children or those who were stunted at age 1 but not at age 8 had lower cognitive scores than their counterparts who never experienced growth faltering.^12^ Other analyses of the Young Lives Study data also reported that linear growth across childhood or in adolescence was associated with cognitive outcomes at ages 12 and 15 years, respectively.^13,14^ In a cohort of Thai children, linear growth from birth to 4 months and from 4 months to 12 months were both positively associated with intelligence quotient (IQ) at 9 years of age.^15^ In a cohort study of Chinese children, weight gain between 6 and 12 months after birth showed positive associations with IQ, comprehension, memory, and reasoning at aged 7-9 years.^16^

Fewer studies have investigated the association between adult IQ and growth across childhood. Two studies in Brazilian cohorts showed that growth in early childhood had a positive association with IQ, school attainment and monthly income in adults.17,18 Given the paucity of studies on this topic and the fact that LMICs have the highest burden of growth faltering in childhood, the long-term correlates of child growth especially in LMICs, need further research.

Although the importance of growth during the first 1,000 days to acquisition of human capital is undisputed,^4-8^ it has been claimed that there is a second window of opportunity during adolescence when catch-up in height may occur.^2,14,19^ In a World Bank Group book about child and adolescent health and development that combines economic theory with health sciences, Alderman et al. agree on the need for a better comprehension of *“individuals’ development timing and age-dependent responses”*.^20^ Therefore, studies are needed to assess how adult human capital is associated with growth during different age ranges from birth to adulthood. This evidence may contribute to better targeted interventions among young children, adolescents, or both groups.

We describe associations between growth across childhood and adolescence and two dimensions of adult human capital (school attainment and IQ) in six birth cohorts from five LMICs.

## Methods

### Study design and data sources

We analyzed data from the six birth cohorts that constitute the Consortium of Health-Oriented Research in Transitioning Societies (COHORTS).^21^ The cohorts are from Brazil,^22,23^ Guatemala,^24^ India, ^25^ the Philippines,^26^ and South Africa .^27^ All fieldwork followed procedures approved by local Ethics Review committees, and the present analyses were approved by the Emory University Institutional Review Board.

### Child growth

Anthropometrics in childhood were obtained using site-specific protocols as described elsewhere.^22-27^ Birth data were collected in hospitals after delivery (Brazil and South Africa), in the community within 3 days of birth by the research team (India), at home or in hospitals by birth attendants (Philippines), and in a healthcare center or at home by a project nurse 15 days after birth (Guatemala). Birth weight was available in all cohorts, and birth length was available in all cohorts except Brazil (1982) and South Africa. Calculation of gestational age was based on the date of last menstrual period reported by the child’s mother, and supplemented with Ballard scores for LBW infants in the Philippines. We identified ages at measurement common across the birth cohorts. Supine length and weight were measured at age 24 months in all cohorts except Brazil (1993), for which measurements were obtained at 12 months. We refer to this age as at around 2 years. Standing height and weight were measured at 48 months for all cohorts (supine length was measured in Guatemala) except for the Philippines, where measures were obtained at 108 months. We refer to this age as mid-childhood (MC). We refer to all postnatal measures of length and height as height, for convenience. Heights and weights were expressed as height-for-age (HAZ) and weight-for-age (WAZ) Z-scores using the WHO Growth Standards.^28^

Repeated measures of height and weight are highly correlated. As in previous work,^5^ we created conditional height measures by regressing current height on all previous height and weight measurements, and conditional weight measures by regressing current weight on current height and all prior height and weight measures, within strata of site and sex. Conditional size variables are standardized (mean=0 and standard deviation=1) residuals of such regressions, and denote how much a child deviates from his/her expected height or weight based on his/her earlier growth, considering the growth trajectories of the other children of the same sex and cohort.

We generated three conditional height variables: conditional height at 2 years, and conditional height in MC and in adulthood. These variables correspond to linear growth from birth to 2 years, from 2 years to MC, and from MC to adulthood, respectively. Regarding weight, three conditional variables were also generated: conditional weight at 2 years, in MC and in adulthood. These variables correspond to relative weight from birth to 2 years, from 2 years to MC, and from MC to adulthood, respectively.

As birth length was not available for two cohorts, we generated conditional measures using birth weight as the anchor. Additionally, results using birth length as the anchor (for the four cohorts for which this measure was available) are provided in the Supplementary tables. Previous analyses have shown that models starting with birthweight or birth length produce similar associations with later outcomes.^5^

### Adult outcomes

We obtained the highest grade of attained formal schooling by interview. We modeled attained schooling as an integer variable. For India, the data are only available in categorized form, and we assigned numeric values based on the years typically required to attain each category. We also categorized the number of years of schooling attainment as a binary variable, using site-specific thresholds relevant to schooling completion current at the time the cohort members were children (Brazil 1982: ≥ 12 years; Brazil 1993: ≥ 11 years; Guatemala: ≥ 6 years; India: ≥ 13.5 years; The Philippines: ≥ 11 years; South Africa: ≥ 12 years).

To measure IQ in adulthood, we administered the Raven’s Standard Progressive Matrices^29^ to participants in the Guatemala, Philippines and South Africa cohorts. In Guatemala, sections A through C were administered due to inability to proceed beyond this point, for a maximum score of 36 points. In the Philippines and South Africa, sections A through E were administered, for a maximum score of 60 points. In both Brazilian cohorts, the arithmetic, digit symbol, similarities, and picture completion subtests of the Wechsler Adult Intelligence Scale (3rd version) were administered.^30^ Adult IQ was not available for the India cohort. We standardized the distribution within each cohort and by sex to a mean of 100 and a standard deviation of 15 to remove between-cohort differences that may relate to language of administration, context, or tests administered

### Covariates

Maternal height (cm), age at birth of the cohort participant (years), schooling (years), paternal schooling (years), child birth order, household socioeconomic status (quintiles of the site-specific distribution), and for Guatemala birth year and intervention group (Atole, Fresco), in the original nutrition supplementation trial near the time of the cohort participant’s birth were extracted from data archives.

### Statistical analysis

We used R version 3.6.2 for analyses. We restricted all analyses to participants with complete information for anthropometric variables at birth and in childhood, and IQ (except India) and schooling variables in adulthood. For descriptive analyses, we calculated means and standard deviations for continuous variables, and proportions for categorical variables.

In cohort-specific analyses, we used linear regression for continuous outcomes (number of years of attained schooling and IQ scores) and quasi-Poisson regression for the binary outcome of school attainment to estimate the associations with conditional size measures. We used inverse sampling weights in all analyses of the 1993 Brazil cohort, because data were collected for all low birthweight infants and a random 20% sample of other infants.

All cohort-specific analyses were performed for males and females separately. We stratified the analyses by study site and sex given observed heterogeneity among sites and previous literature that supports sex differences in IQ scores.^31^ Sex-combined estimates were generated by pooling the sex-specific estimates using weighted random effects meta-analysis, with the weight each sex received being proportional to sample size.

A doubly-robust strategy was used for covariate adjustment, where adjustment was performed via multivariable regression after inverse probability of treatment weighting (IPTW) using the “ipwpoint” function from the “ipw” package in R,^32^ applying linear regression to model the relationship between the exposure variable and the covariates. This regression was specified to include a “main effect” term for all covariates, as well as all pairwise product terms between covariates. To mitigate the possibility that individuals with large weights could substantially influence the results, the left tail of the weights was truncated at the 0.5th percentile, and the right tail at the 99.5th percentile.

We defined four models a priori with progressive adjustment of potential confounders. We used listwise deletion in all models. In model one (minimal adjustment), we adjusted for sex (for analyses involving both sexes), and year of birth and intervention group in Guatemala. In model two, we controlled for the same variables as in model one but excluded cases that could not be included in subsequent models because of missing values. We found similar point estimates between models one and two, suggesting that missing covariate data was not a major factor in our results. In model three, we adjusted for the covariates in model one, plus early-life socioeconomic quintiles, maternal schooling, maternal age, maternal height, birth order and, for the Brazil 1982 and 1993 cohorts, skin color. Comparing models two and three allowed us to assess changes due to confounding by the selected covariates. In model four (further adjustment), we controlled for covariates in model three plus paternal schooling. Model four is our preferred representation.

We used random effects meta-analysis to pool the sex- and cohort-specific results. Pooled sex-combined estimates were generated by pooling the corresponding pooled sex-specific estimates. The variation between cohorts was estimated using *I*^2^ statistic and Cochran’s Q test, and random effects meta-regression was used to test for the effect modification by sex.

## Results

Data from 9503 participants with complete data for at least one of the outcomes and size at 2 years of age were analyzed. Table 1 shows selected characteristics of the participants. Birth weight z-scores were lowest among individuals born in India. The Guatemalan, Filipino and Indian participants had the lowest height-for-age and weight-for-age z-scores in early and mid-childhood and were shortest as adults. Schooling for both parents was lowest in Guatemala. School attainment was higher in women across all cohorts when compared to men, except in Guatemala.

**Table 1.** Characteristics of participants in the six cohorts stratified by sex.

|  | Brazil<br>1982 |  | Brazil<br>1993 |  | Guatemala<br>1969-77 |  | India<br>1969-72 |  | Philippines<br>1983-84 |  | South Africa<br>1990 |  |
| --- | --- | --- | --- | --- | --- | --- | --- | --- | --- | --- | --- | --- |
|  | Men<br>(n=1606) | Women<br>(n=1657) | Men<br>(n=564) | Women<br>(n=629) | Men<br>(n=332) | Women<br>(n=394) | Men<br>(n=840) | Women<br>(n=614) | Men<br>(n=901) | Women<br>(n=813) | Men<br>(n=548) | Women<br>(n=605) |
| **Birth variables** |  |  |  |  |  |  |  |  |  |  |  |  |
| Gestational age (weeks) | 39.4 (1.7) | 39.4 (1.8) | 39.2 (2.6) | 39.1 (2.8) | 39.1 (2.8) | 39.6 (3.1) | 38.7 (2.6) | 39.1 (2.4) | 38.7 (2.2) | 38.8 (2.1) | 38.1 (1.8) | 38.0 (1.9) |
| Birth length (WHO Z-scores) | NA | NA | -0.8 (1.6) | -1.0 (1.5) | 0.0 (1.3) | 0.0 (1.1) | -0.7 (1.1) | -0.6 (1.1) | -0.3 (1.1) | -0.2 (1.1) | NA | NA |
| Birth weight (WHO Z-scores) | -0.2 (1.1) | -0.2 (1.2) | -0.7 (1.5) | -0.9 (1.5) | -0.7 (1.1) | -0.6 (1.0) | -1.1 (1.0) | -1.1 (0.9) | -0.7 (1.0) | -0.6 (1.0) | -0.6 (1.1) | -0.6 (1.2) |
| **Childhood variables** |  |  |  |  |  |  |  |  |  |  |  |  |
| HAZ at around 2 years (SD) | -0.7 (1.2) | -0.6 (1.2) | -0.3 (1.3) | -0.2 (1.2) | -3.0 (1.1) | -2.9 (1.1) | -2.0 (1.2) | -1.9 (1.1) | -2.4 (1.1) | -2.4 (1.1) | -1.4 (1.1) | -1.1 (1.0) |
| HAZ in mid-childhood (SD) | -0.6 (1.1) | -0.6 (1.1) | -0.2 (1.1) | -0.2 (1.1) | -2.4 (0.9) | -2.5 (1.0) | -1.9 (1.0) | -2.0 (1.0) | -2.1 (0.9) | -2.0 (0.9) | -1.0 (0.9) | -0.9 (0.9) |
| WAZ at around 2 years (SD) | 0.1 (1.1) | 0.1 (1.0) | 0.4 (1.2) | 0.4 (1.0) | -1.8 (1.0) | -1.7 (1.0) | -1.5 (1.1) | -1.4 (1.1) | -1.7 (1.0) | -1.7 (1.0) | -0.7 (1.1) | -0.3 (1.0) |
| WAZ in mid-childhood (SD) | 0.1 (1.0) | 0.0 (1.0) | 0.3 (1.2) | 0.2 (1.1) | -1.4 (0.8) | -1.5 (0.9) | -1.3 (0.9) | -1.5 (0.9) | -2.0 (1.0) | -1.9 (0.9) | -0.5 (0.9) | -0.5 (0.9) |
| **Adult variables** |  |  |  |  |  |  |  |  |  |  |  |  |
| Age (years) | 30.2 (0.3) | 30.2 (0.3) | 22.6 (0.3) | 18.4 (0.3) | 46.5 (2.6) | 46.2 (2.6) | 36.1 (1.1) | 36.1 (1.0) | 34.4 (0.5) | 34.5 (0.5) | 28.5 (0.4) | 28.5 (0.4) |
| Height (cm) | 173.8 (6.9) | 161.1 (6.2) | 174.1 (7.7) | 160.2 (6.8) | 163.7 (5.8) | 151.1 (5.7) | 169.6 (6.4) | 154.9 (5.6) | 162.9 (5.9) | 151.1 (5.4) | 171.5 (6.4) | 159.5 (6.2) |
| BMI (kg/m<sup>2</sup>) | 27.2 (5.1) | 26.8 (6.0) | 24.6 (4.7) | 25.4 (5.8) | 26.7 (4.3) | 29.4 (5.1) | 26.9 (4.6) | 27.5 (5.2) | 24.8 (4.4) | 25.2 (5.0) | 21.6 (3.9) | 25.8 (6.1) |
| **Covariates** |  |  |  |  |  |  |  |  |  |  |  |  |

|  | **Brazil**<br>**1982** |  | **Brazil**<br>**1993** |  | **Guatemala**<br>**1969-77** |  | **India**<br>**1969-72** |  | **Philippines**<br>**1983-84** |  | **South Africa**<br>**1990** |  |
| --- | --- | --- | --- | --- | --- | --- | --- | --- | --- | --- | --- | --- |
|  | Men<br>(n=1606) | Women<br>(n=1657) | Men<br>(n=564) | Women<br>(n=629) | Men<br>(n=332) | Women<br>(n=394) | Men<br>(n=840) | Women<br>(n=614) | Men<br>(n=901) | Women<br>(n=813) | Men<br>(n=548) | Women<br>(n=605) |
| Maternal age at childbirth (years) | 26.1 (6.2) | 26.2 (6.4) | 26.3 (6.6) | 26.3 (6.2) | 27.1 (7.2) | 27.3 (7.1) | 26.9 (6.0) | 26.9 (5.6) | 26.4 (6.1) | 26.4 (6.0) | 25.6 (6.3) | 25.7 (6.3) |
| Maternal height (cm) | 156.6 (6.3) | 156.5 (5.8) | 159.7 (6.6) | 159.7 (6.7) | 148.1 (4.8) | 148.3 (5.1) | 151.8 (5.6) | 152.0 (5.1) | 150.5 (4.9) | 150.5 (5.0) | 158.2 (5.8) | 158.7 (5.8) |
| Maternal schooling (years) | 6.5 (4.0) | 6.7 (4.2) | 6.7 (3.5) | 6.5 (3.4) | 1.3 (1.5) | 1.3 (1.5) | 5.5 (4.5) | 5.7 (4.5) | 7.0 (3.3) | 6.8 (3.1) | 9.7 (2.4) | 9.6 (2.6) |
| Paternal schooling (years) | 6.7 (4.1) | 6.9 (4.2) | 6.6 (3.5) | 6.9 (4.2) | 1.8 (2.2) | 1.8 (2.1) | 10.4 (5.1) | 11.2 (4.7) | 7.2 (3.5) | 6.9 (3.3) | 10.6 (2.4) | 10.5 (2.5) |
| Birth order |  |  |  |  |  |  |  |  |  |  |  |  |
| 1 | 40.6% | 38.9% | 35.6% | 33.1% | 18.5% | 14.0% | 16.4% | 14.0% | 21.8% | 21.8% | 37.4% | 38.7% |
| 2 | 28.7% | 29.0% | 24.5% | 28.5% | 12.7% | 14.7% | 23.1% | 20.9% | 21.6% | 23.0% | 29.2% | 28.4% |
| 3 | 16.0% | 16.0% | 19.1% | 22.1% | 12.1% | 15.0% | 20.7% | 21.4% | 19.6% | 18.9% | 17.0% | 17.4% |
| ≥4 | 14.7% | 16.1% | 20.7% | 16.4% | 56.7% | 56.3% | 39.8% | 43.8% | 37.0% | 36.3% | 16.4% | 15.5% |
| **Human capital variables** |  |  |  |  |  |  |  |  |  |  |  |  |
| IQ score (standardized ) | 100.8 (15.2) | 99.7 (14.7) | 99.1 (15.9) | 99.6 (14.1) | 105.8 (15.6) | 97.8 (13.4) | NA | NA | 100.3 (14.8) | 99.6 (15.2) | 100.6 (14.4) | 99.4 (14.8) |
| School attainment (years of schooling) | 10.9 (4.0) | 11.9 (4.2) | 9.2 (2.7) | 10.0 (2.3) | 5.6 (3.7) | 4.7 (3.7) | 13.2 (3.4) | 13.9 (3.1) | 10.2 (3.4) | 11.2 (2.9) | 11.5 (1.6) | 11.9 (1.4) |

Data are presented as mean (SD) or percentage. NA=not available data. HAZ= height-for-age Z-score. WAZ=weight-for-age Z-score. IQ=intelligence quotient. Sample sizes refer to participants with non-missing data for at least one outcome and non-missing data for either HAZ or WAZ at 2 years of age. Adult age was estimated based on date of IQ measurement, except India (age at latest wave of data collection).

### Growth and school attainment

The site- and sex-pooled associations between growth and school attainment are presented in Table 2 and Supplementary Table 2. Measures of weight or length at birth, and height at around 2 years and in MC were each positively associated (p<0.01) with schooling attainment. Covariate adjustment attenuated the estimates and linear growth in MC was no longer significant. The point estimates were larger for conditional height at 2 years of age (0.25, 95%CI: 0.10, 0.40) compared to birth weight (0.13, 95%CI: 0.08, 0.19) or birth length (0.20, 95%CI: 0.11, 0.30). Conditional height in adulthood was not associated with schooling attainment in men (p=0.48) but it was inversely associated in women (p=0.013). Conditional relative weight at around 2 years was not associated with schooling attainment in fully adjusted models. Conditional relative weight in MC and in adulthood was inversely associated with schooling attainment. The results were generally consistent between males and females and across the six cohorts, although there was heterogeneity in the size of the estimates (Table 3). Sex differences were observed in the estimates for conditional height in adulthood, and for the estimates for conditional relative weight in MC and in adulthood. In models that examined the binary categorization of schooling attainment, we found similar associations in both pooled and site-stratified analyses (Supplementary Tables 3 and 5).

**Table 2.**
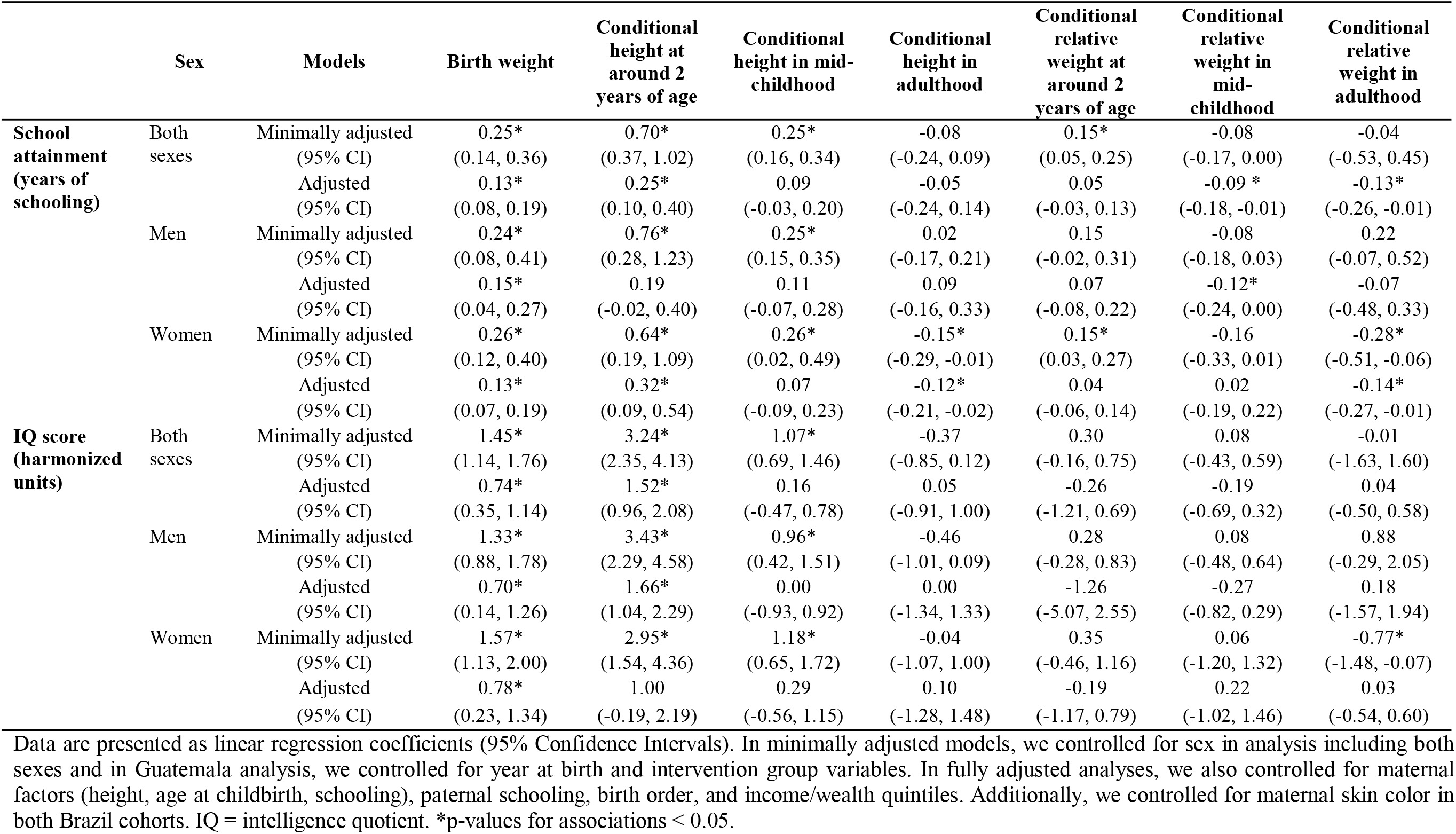
Pooled adjusted associations between growth in childhood and school attainment and IQ in adulthood, by sex.

**Table 3.**
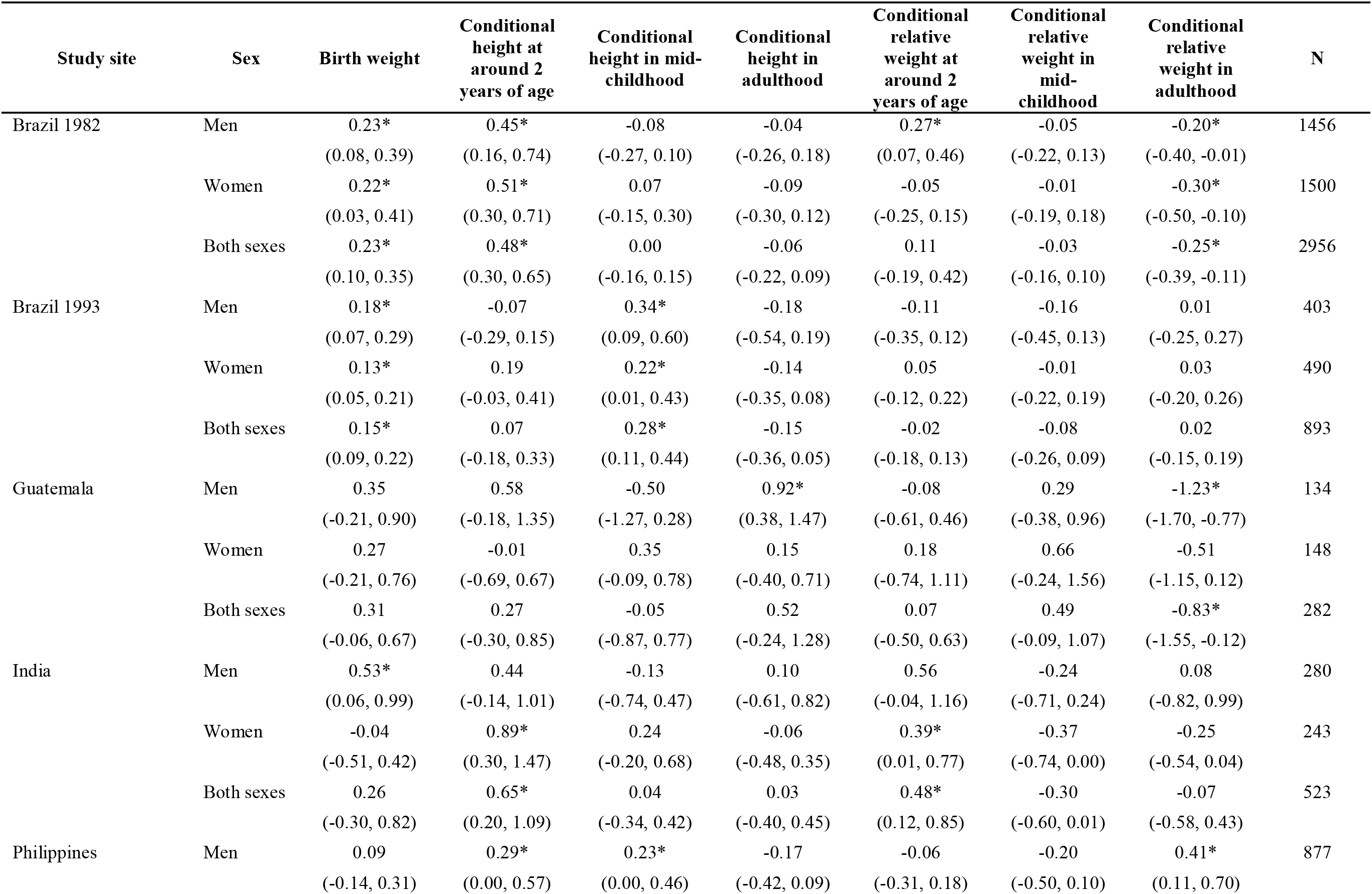
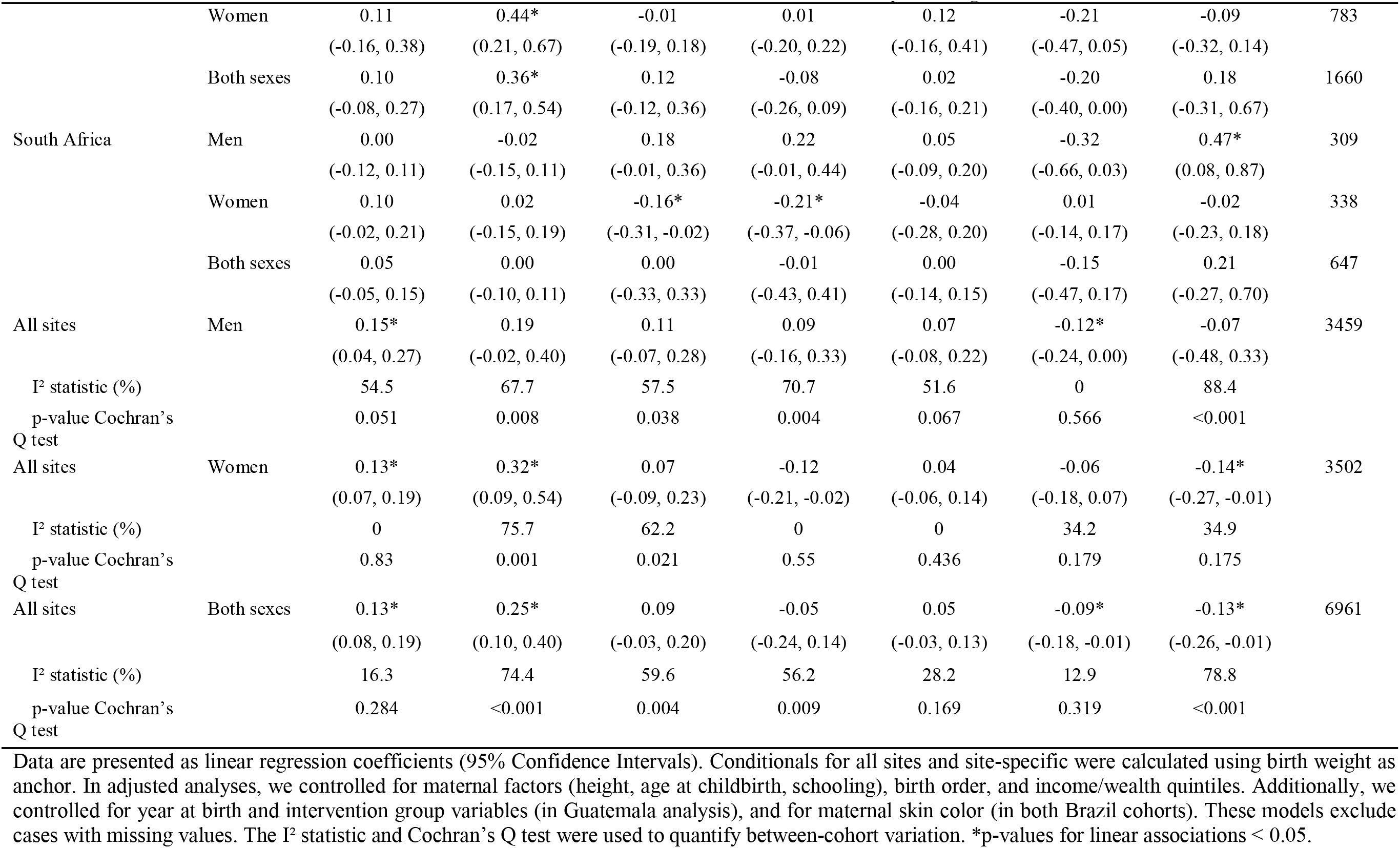
Adjusted associations between growth in childhood and the number of years of school attainment in adulthood, by study site and sex.

### Growth and IQ

The site- and sex-pooled associations between growth and IQ are presented in Table 2 and Supplementary Table 2. Measures of weight or length at birth, and height at around 2 years and in MC were each positively associated (p<0.001) with adult IQ. After covariate adjustment the strength of associations was reduced and the estimate for conditional height in MC was not significant. The point estimates were larger for conditional height at 2 years of age (1.52, 95%CI: 0.96, 2.08) compared to birth weight (0.74, 95%CI: 0.35, 1.14) or birth length (0.73, 95%CI: 0.35, 1.10). Conditional height in adulthood was not associated with IQ. Measures of relative weight at around 2 years, in MC and in adulthood were not associated with adult IQ. In general, the results were consistent between males and females although there was heterogeneity across the five cohorts in the size and significance of the estimates (Table 4).

**Table 4.**
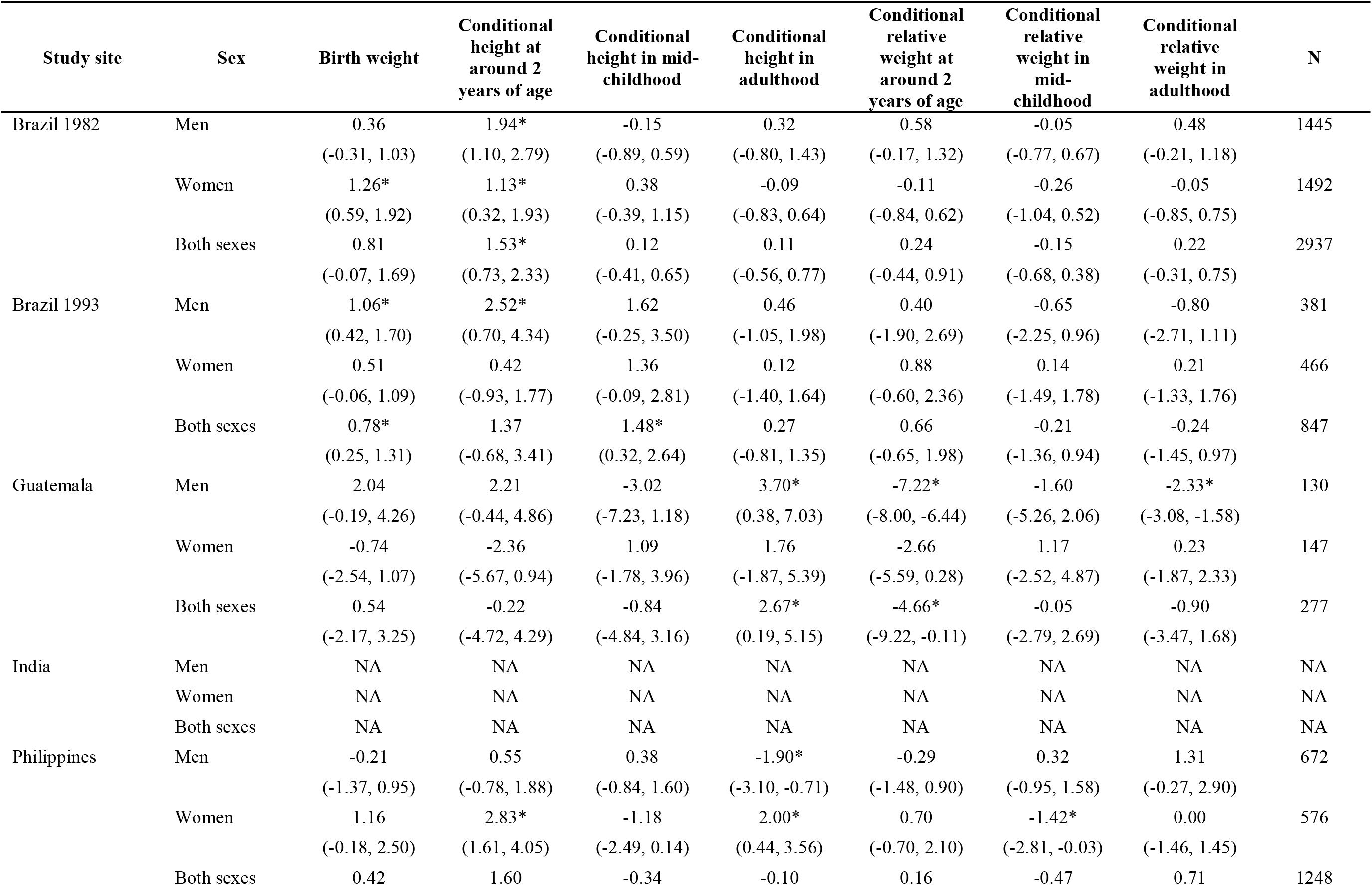
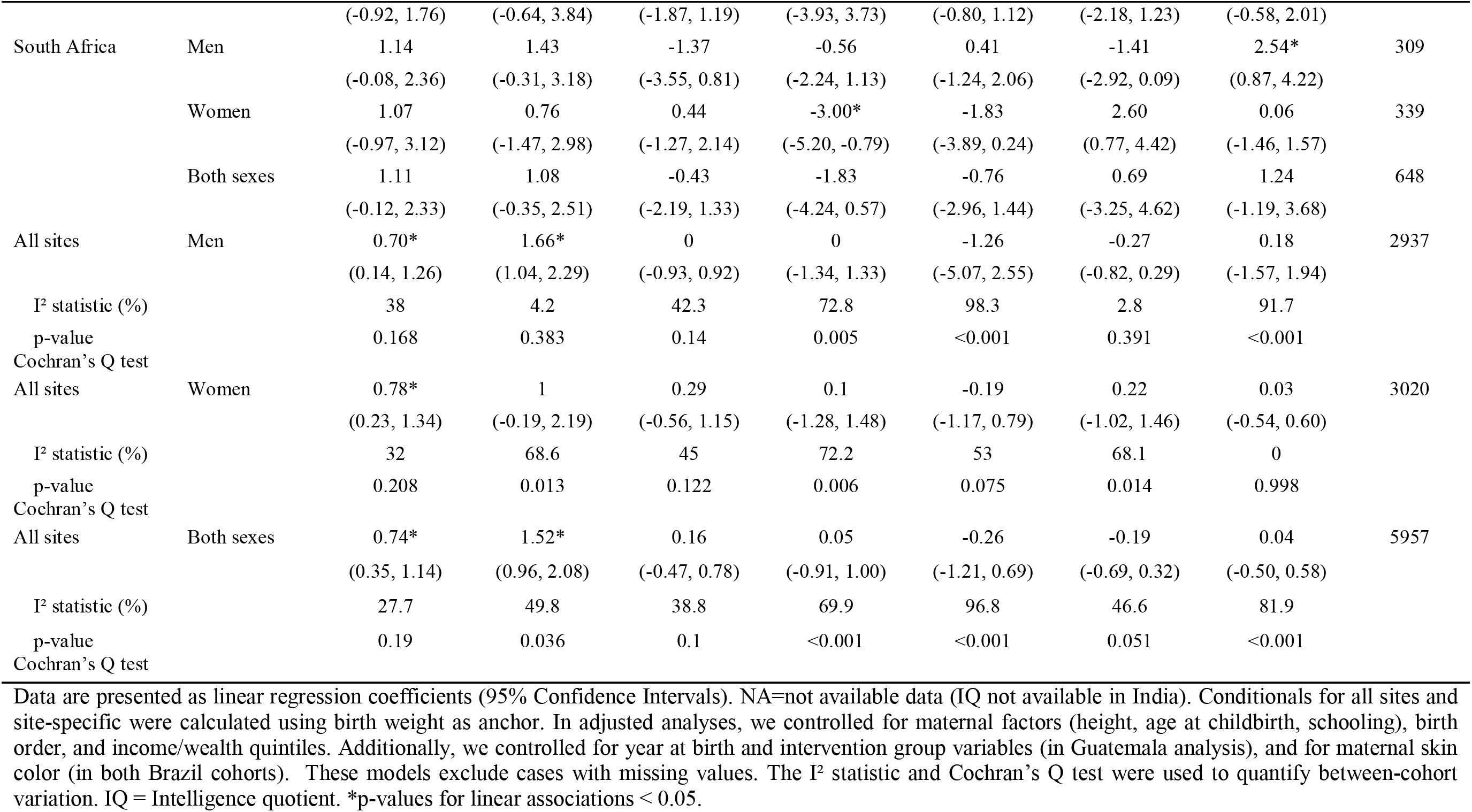
Adjusted associations between growth in childhood and IQ in adulthood, by study site and sex.

## Discussion

This analysis of data from six birth cohorts from LMICs showed that birth size and linear growth from birth to age 2 years were positively associated with schooling attainment and adult IQ. Linear growth from age 2 years to MC was not associated with either of the outcomes in fully adjusted models. We observed that the effect sizes were larger for conditional height at 2 years of age when compared to length at birth. Apart from an inverse association between conditional height in adulthood and schooling attainment in female participants, conditional height in adulthood was not associated with either schooling attainment or IQ. Conditional relative weight in MC and in adulthood were each inversely associated with schooling attainment. Change in relative weight was not associated with adult IQ for any of the age intervals examined.

These results suggest that birth size and linear growth from birth to around 2 years are independent predictors of schooling and intelligence in adulthood. This is important because improvements in human capital dimensions are associated with economic growth. This independent association, at least in childhood, had previously been observed.^12,13,33,34^ We found that for every 1 Z-score increase in conditional linear growth at age 2 years there was an increase in 0.25 years of schooling and 1.52 units of adult IQ score after adjustment (Table 2). It has been estimated that for every additional year of schooling there is a 7.9% country level economic return;9 and one standard deviation increase in cognitive skills of a country’s workforce has been associated with an increase of two percentage points in the per capita GDP (annual growth) and economic returns that range from 0.07 to 0.48, in developing countries.^35,36^

Our findings further confirm that timing (and specifically the first 1,000 days) is critical to improve schooling outcomes and intellectual performance in adulthood. Human growth is not uniform and systems, organs, and tissues develop at different velocities. Linear growth is characterized by high initial velocity with rapid deceleration in the first two years after birth.^37^ The brain achieves 83% of its adult volume by 24 months of postnatal life.^38^ Thus, shared underlying determinants of both length and neurodevelopment might influence adult human capital defined as a group of capacities, abilities, and intangible assets useful to create economic value.^39^ Interventions in early life have the highest overall returns compared to other life course stages, and interventions to reduce stunting in the first 24 months of age not only increased preschool linear growth but had benefit-cost ratios greater than one, demonstrating that such interventions are a good economic investment.^20^ In that sense, that benefit in linear growth also indirectly benefit schooling attainment and cognitive performance in the long term.

We infer that the multiple causes of linear growth failure partly impact on brain development and learning; hence, linear growth failure predicts schooling and IQ. The relationship is not causal and there are at least four pathways that explain the results observed. First, linear growth and brain development are susceptible to common nutritional inputs,^40^ many of which have underlying determinants such as socioeconomic status (poverty), maternal education, food insecurity, water scarcity, poor sanitation and hygiene, among others.^41^ Thus, nutritional deficits in sensitive periods of life affect not only body growth but brain size, structure, development and function.^42^ Second, growth faltering makes children more susceptible to infections that in turn decrease appetite, absorption, and nutrients’ use; nutrients will be diverted to the immune system affecting its availability for growth and development.^43^ Third, undernourished and ill children are commonly apathetic, irritable, and less interested in exploring their environment.^43^ Fourth, the size of children may elicit different interactions with adults given that short children appear younger and are treated as such.^44^ Thus, short children lag in acquiring motor, cognitive and social behaviors.^44^

Our results confirm earlier research that prenatal growth and linear growth in early childhood are significantly associated with higher school attainment^4-7,17,18^ and adult IQ.^7,17,18^ Analyses of Young Lives Study data found that linear growth in different age intervals (1, 5, 8, 12, and 15) was positively associated with cognitive skills in mid-childhood, early adolescence and adolescence.^12-14,33^ Two studies suggested that these associations were mediated through HAZ at subsequent age intervals.^13,33^ We observed similar tendencies in the direction of the associations, however, the magnitudes of our estimates were stronger than those reported in the Young Lives Study.^12,14,33^

We did not find statistical associations between conditional relative weight in early childhood and school attainment after full adjustment. Adair et al^5^ had previously observed a significant but weak association between conditional relative weight in early childhood and the highest grade attained. The differences in our study might be attributed to the inclusion of a new birth cohort (Brazil 1993), variations in sample sizes, and a higher number of covariates. Conditional relative weight in early and in MC were not associated with IQ, as previously shown in studies conducted with adults^17,18^ and children.^15^

Our findings agreed with previous studies that linear growth from 2 years to MC and from MC to adulthood were not associated with schooling attainment and adult IQ.^5,17,18^ Indeed, we found that conditional height and change in relative weight from MC to adulthood were inversely associated with schooling attainment in female participants. Our results do not support the notion that interventions in adolescence aimed at improving linear growth are likely to impact adult IQ, as has been suggested by other authors.^14^ The period from MC to adulthood includes late childhood, adolescence and adulthood through ages 18 to 46 years. Linear growth and neurodevelopment are largely complete by adulthood and individuals have already finished school. Thus, it is unlikely that weight changes in adulthood are predictors of schooling attainment. Rather, the inverse associations may reflect reverse causality. In adulthood, women’s weight and weight changes are influenced by multiple biological and environmental factors such as genetics (body composition), reproductive events (parity), marital status, life-style behaviors, work settings, food environment, socioeconomic status, many of which might be determined schooling attainment earlier in life.

Among the study’s limitations, we note some inconsistencies in the ages of exposures and outcomes across the cohorts. Height was measured at 2 years of age in all sites except Brazil 1993 cohort, where it was measured at 1 year of age. Similarly, MC was considered to be 4 years in most of the cohorts, but 8.5 years in the Philippines. There were also differences in the ages in which the schooling and IQ outcomes were obtained, and some differences in the instruments used to measure IQ across sites. Finally, residual and unmeasured confounding should be considered given the observational nature of our study.

This study has several strengths. We analyzed six well-characterized population-based birth cohorts in five LMIC with follow-up periods that ranged from 18 to 46 years. Each study site had trained staff who followed standard methodologies to collect the anthropometric and sociodemographic data, which minimized measurement error and recall bias. The treatment of our exposures as conditional measures of growth avoids collinearity and facilitates differentiation of linear growth at specific age intervals. Each site used validated measures of intelligence, and we standardized the distributions within each cohort and by sex to be able to compare outcomes. We adjusted our models for a range of early life social factors. Additionally, our analytic sample with complete data was not affected by missing values and loss at follow-up, as evidenced by the similar results obtained when comparing models one and two (Supplementary Table 1). We saw evidence of heterogeneity among the cohorts in the magnitude of the estimates but not in the direction of the associations. Thus, we were able to obtain single pooled estimates combining cohort-specific results. These associations might be generalized to other LMICs, but should be interpreted with caution given the differences between pooled and site-specific results, acknowledging that our findings might not represent the entire diversity and complexity of the studied countries.

In conclusion, our results show an independent association between prenatal growth and linear growth from birth until the second year of life with adult human capital. These findings confirm the importance of the first 1,000 days, a sensitive period where adversities including poor nutrition will have long-lasting effects on adult size and functional capacities such as learning and intelligence. Our findings do not show that improvements in linear growth after the first two years, during childhood, adolescence or adulthood, were associated with adult human capital outcomes.

## Data Availability

The data is available under reasonable request.

## Contributors

ADS and CGV conceived the idea and study design. NEP performed the literature review and wrote the first draft of the manuscript. NEP, ADS, CGV and FPH led the writing team. FPH performed the statistical analyses. CGV, LSA, FCB, SKB, BLH, NRL, RM, MM, AMBM, SAN, LMR, HSS, AS, FCW, and ADS participated in data collection. All authors read and commented on successive drafts, and approved the final manuscript.

## Acknowledgments

We acknowledge funding from the Bill and Melinda Gates Foundation (OPP1164115) for data collection in Guatemala, Philippines and South Africa and for data management and analysis. Data collection in Brazil was funded by the Wellcome Trust. The New Delhi Birth Cohort has received funding from the Indian Council of Medical Research, the Department of Biotechnology, the United States National Center for Health Statistics, the Medical Research Council (UK), the British Heart Foundation, and the Bill and Melinda Gates Foundation. The Birth to Twenty Plus Cohort is supported by the South African Medical Research Council, DSI-NRF Centre of Excellence in Human Development at the University of the Witwatersrand, Johannesburg, South Africa, and the Wellcome Trust (UK).

